# RT-qPCR assay for detection of British (B.1.1.7) and South Africa (B.1.351) variants of SARS-CoV-2

**DOI:** 10.1101/2021.02.25.21252454

**Authors:** Karin Yaniv, Eden Ozer, Noam Plotkin, Nikhil Suresh Bhandarkar, Ariel Kushmaro

## Abstract

Less than a year following the SARS-CoV-2 outbreak, variants of concern have emerged in the form of the British variant B.1.1.7 and the South Africa variant B.1.351. Due to their high infectivity and morbidity, it is crucial to quickly and effectively detect them. Current methods of detection are either time-consuming, expensive or indirect. Here, we report the development of a rapid, cost-effective and direct RT-qPCR method for detection of the two variants of concern. We developed and validate a detection system for the detection of the B.1.1.7 variant and another single detection set for the B.1.351 variant. The developed approach was characterized and tested on wastewater samples and illustrated that all primers and probes were sensitive and specific. The novel system presented here will allow proper response and pandemic containment with regard to these variants. In addition, it may provide a basis for developing tools for the detection of additional variants of concern.

## Introduction

The SARS-CoV-2 world pandemic erupted early 2020 with rising numbers in morbidity and mortality. SARS-CoV-2 was recognized as an RNA virus, therefore detection methods emerging for immediate response were mainly in the form of reverse transcriptase quantitative polymerase chain reaction (RT-qPCR) (Lu et al., 2020). In RT-qPCR, RNA is extracted, undergoes reverse transcription for DNA strand generation, followed by PCR amplification and TaqMan probes fluorescence detection. To date, RT-qPCR is the most common methodology for SARS-CoV-2 diagnostics (Vogels et al., 2020).

Starting in September 2020, new variants of concern of SARS-CoV-2 virus began to emerge. Amongst them, the variants termed the British variant B.1.1.7 and South Africa (SA) variant B.1.351 became dominant compared to the original SARS-CoV-2 virus (Wang et al., 2021). Due to their higher infection rate and high morbidity, identification of these variants became essential. A diagnostic tool that could quickly and efficiently distinguish between the variants is imperative to help evaluate the variants’ distribution. Proper “variant mapping” will provide much needed information to enforce appropriate policy for pandemic containment.

Currently, three methodologies have been developed for SARS-CoV-2 variant diagnostics. The first methodology, that is still mainly being employed, is the next generation sequencing (NGS) approach (Andrés et al., 2020; Khan et al., 2020). In NGS the entire variant’s genome is sequenced. Despite demonstrating the importance of this technique for identification of new variants, its use on previously sequenced variants is time-consuming and requires significant financial means.

Additional detection methods are based on RT-qPCR and include a “drop-out” signal, available in commercial kits (such as TaqPath COVID-19 diagnostic tests, Thermo Scientific, Helix^®^ COVID-19 Test) or a method published in a recent study (Vogels et al., 2021). These use RT-qPCR with two different markers, a double signal manifest for the original SARS-CoV-2 virus, while only a single signal manifest for the targeted variant. Another detection methodology is through characterization of ΔCt between one detection signal and another amongst the different variants (Kovacova et al., 2021). Thus theses current RT-qPCR approaches for variant detection, though significantly faster and cost-effective than the NGS methodology, focuses on indirect detection and may result in false/inconclusive identification.

Therefor there is still a great need for a quick improved specificity- and sensitivity SARS-CoV-2 variants detection methods. Such advanced methodologies will be amenable for clinical diagnostics, as well as for environmental-derived quantification, greatly improving wastewater and population level epidemiology. In this study, we developed a RT-qPCR assay for the direct detection of SARS-CoV-2 British variant B.1.1.7 and another set of RT-qPCR primers-probe for the detection of SARS-CoV-2 SA variant B.1.351. Our design was tested on S gene deletion and non-deletion DNA templates and RNA originating from wastewater samples to assess the sensitivity and specificity of the described sets.

## Methods

### Primers and probes design

The original sequence of SARS-CoV-2 (NC_045512.2) was taken from NCBI database. British B.1.1.7 variant (EPI_ISL_742238) and SA B.1.351 variant (EPI_ISL_736935) sequences were taken from GISAID database (Shu and McCauley, 2017). The probe design focused on the S gene 21724-21828 bp location that includes the British deletion 69-70 or S gene 22243-22331 bp location that includes the SA deletion 241-243. All primers and probes were purchased through Integrated DNA Technologies (IDT). ZEN Quencher was added to the probes as a second, internal quencher in qPCR 5’-nuclease assay. To allow a possibility for duplex assay, S1 probe was assigned a 6-carboxy-fluorescein (FAM) fluorophore and SΔ69 probe was assigned to Yakima Yellow (YakYel) fluorophore. SΔ241 probe was assigned with FAM as well.

### RT-qPCR

RT-qPCR was executed using One Step PrimeScript III RT-qPCR mix using standard manufacture protocol (RR600 TAKARA, Japan). Each reaction mixture contains primers (0.5 µM each), probe (0.2 µM each), ROX reference dye and 5 μL of DNA or RNA (dH2O was added to a final volume of 20 µL reaction volume). RT-qPCR amplification was executed using Step One Plus real-time PCR system (Applied Biosystems, Thermo Scientific). In addition to what is described above, in each run all RT-qPCR experiments included quality controls. The first control was using water sample instead of DNA/RNA (Non template control (NTC)). The second control, used for RNA extractions, was MS2 phage detection (Dreier et al., 2005).

### Calibration curves and limit of detection determination

Calibration curves were performed on a known-positive DNA gene block. Two different gene blocks were used; one containing SARS-CoV-2 S gene sequence as reported for Wuhan-Hu-1 (NC_045512.2), the second containing S gene sequence matching the reported 69-70 deletion of the British variant (B.1.1.7) as well as the reported 241-243 deletion of the SA variant (B.1.351). Calibration of S1 probe was performed using S gene sequence from NC_045512.2, while calibration of SΔ69 probe and SΔ241 was performed using S gene sequence with the relevant deletions. Serial dilutions for the relevant gene block were prepared based on copy number calculations. The resulted Ct values were plotted against the log copy number of the S gene template. Linear regression was performed between the log copy number and the Ct values from the RT-qPCR results.

### Wastewater RNA extraction

For wastewater sampling, composite sewage samples from the wastewater treatment plant (WWTP) were immediately transferred to the lab under chilled conditions. The samples were kept at 4°C until processed. Direct RNA was extracted according to manufacture protocol as it describes in NucleoSpin RNA extraction kit (Macherey Nagel, Germany). An amount of 10^5^ copies of the phage was added to the lysis buffer in each RNA extraction for inner control. RNA was eluted with 50 μL of RNase free water and kept at –80°C.

### Complex matrix detection

Extracted RNA from wastewater sample, known to be SARS-CoV-2 negative, was supplemented with known concentrations of a desired gene block. The samples underwent the same RT-qPCR conditions as described for the calibration curves. Results were plotted to represent the new probes limit of detection in a complex environment.

## Results and Discussion

Developing our assay, we focused on the most dominant variants of SARS-CoV-2 currently known. These, the British variant B.1.1.7 and South Africa (SA) variant B.1.351, were deemed the most urgent variants in need for fast detection. Our design for RT-qPCR detection assays of the two variants (Figure 1), is based on the differences in the S gene from the original sequence (NC_045512.2). B.1.1.7 S gene contains a deletion known as Δ69-70 and B.1.351 S gene contains a deletion known as Δ241-243. Accordingly, our designed focused on these regions.

**Figure 1:**
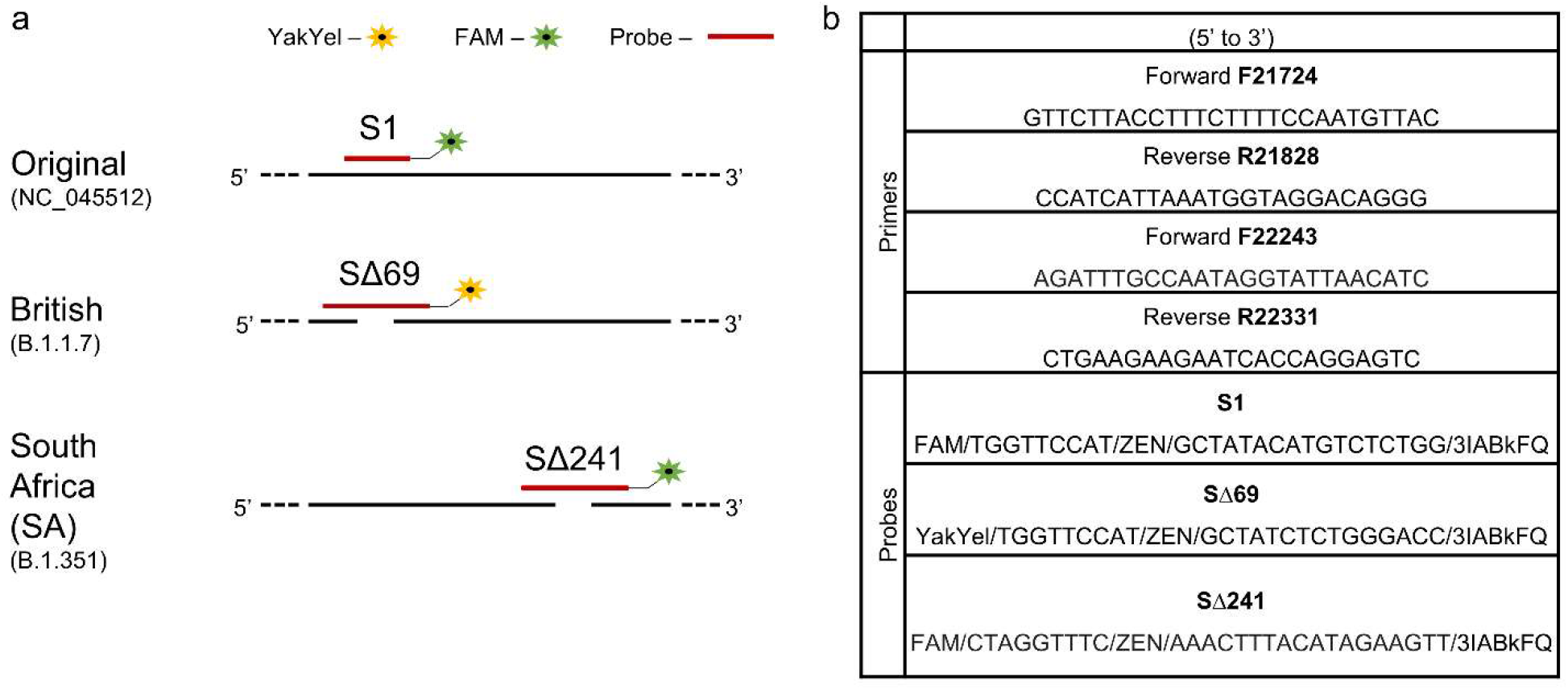
(a) Designed detection set for differentiation and identification of SARS-CoV-2 British variant B.1.1.7 and South Africa variant B.1.351. (b) List of primers and probes sequences.

For B.1.1.7 detection, the designed detection set is located at the S gene 21724-21828 bp of the original sequence. Within this range, the original SARS-CoV-2 and B.1.1.7 sequences are completely identical, apart from 6 nucleotides deletions (Fig. 1a). Our main attempt was to create two separate detections to the amplified area, one corresponding to the original sequence (when using S1 probe) and the other corresponding to the B.1.1.7 (when using the SΔ69 probe). Using designated primers (Fig. 1b) to amplify the specified region surrounding the 6 nucleotides differences, an amplification will be generated regardless to the variant. The probes can thus be used in a single duplex assay via separate wavelengths, where a signal signifies a direct detection of either the original sequence or of B.1.1.7.

For B.1.351 detection, the designated detection region was chosen further along the S gene when compared to the B.1.1.7 detection region. Focusing on S gene 22243-22331 bp of the original sequence, the original SARS-CoV-2 sequence is identical to the B.1.351 sequence apart from a 9 nucleotides deletion (Fig. 1a). Using a detection set comprised of two primers meant to amplify the target region, a single probe (SΔ241 probe) was designed for the detection of the B.1.351 variant. The SΔ241 probe is meant to correspond only to the deletion of 9 nucleotides in the specified region, characterizing B.1.315, therefore will signal detection only when B.1.351 is present and will not correspond to the original sequence.

To ensure functionality, the described sets of primers and probes underwent characterization. Initially, a calibration curve was generated for primers with the relevant probe separately, using dsDNA as a template. A detection range of between 10^7^ copies and ten copies per µL was tested for each probe. Results plotted for both probes confirmed the chosen primers’ (4 different primers, 2 different amplification sets) ability to amplify the target region (Fig. 2). Furthermore, linear regression performed for both probes demonstrated strong coloration and the probes validity for usage on the amplified fragment. A limit of detection (LOD) could be determined for each probe and was identified as 10^1^ copies per µL for all three probes.

**Figure 2:**
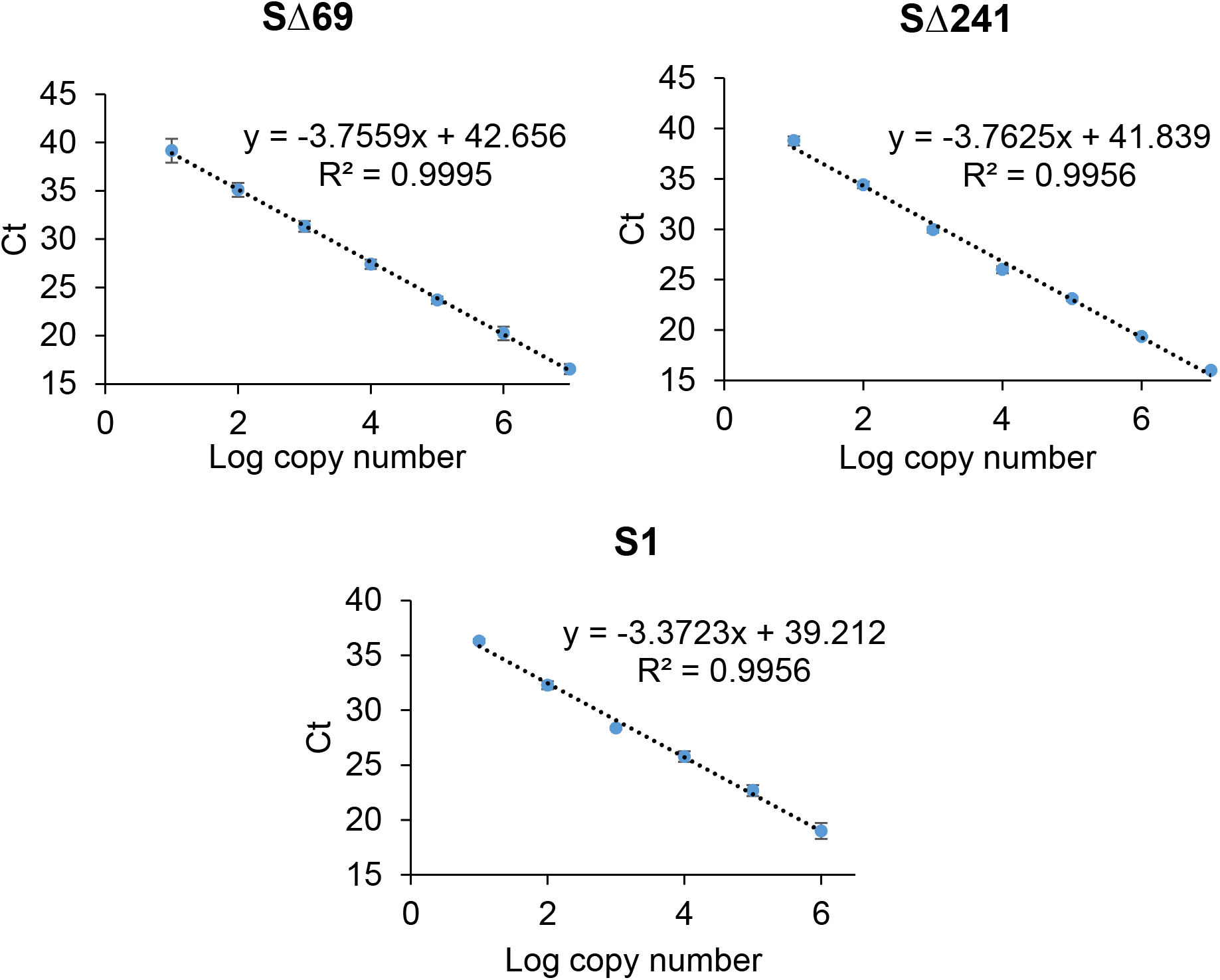
calibration curves and limit of detection of SΔ69 probe, SΔ241 probe and S1 probe.

Following basic characterization, further confirmation to the described methodology was performed using a more complex environment for the RT-qPCR reaction. As SARS-CoV-2 wastewater detection is important with regards to the development of a quick early warning system for virus detection during the global pandemic (Bar-Or et al., 2020), wastewater matrix collected from wastewater treatment plant at the city of Beer Sheva, Israel, was chosen as complex environment. All three probes were employed on wastewater samples pre-determined as negative for SARS-CoV-2 with various dsDNA template copies (Fig. 3). As can be seen in Figure 3, despite the wastewater matrix, the 3 designed probes displayed high detection sensitivity. With the ability to detect up to 10^1^ copies per µL, the new probes demonstrated satisfying detection ability when compared to previously described primers and probes sets for clinical diagnostics (Vogels et al., 2020).

**Figure 3:**
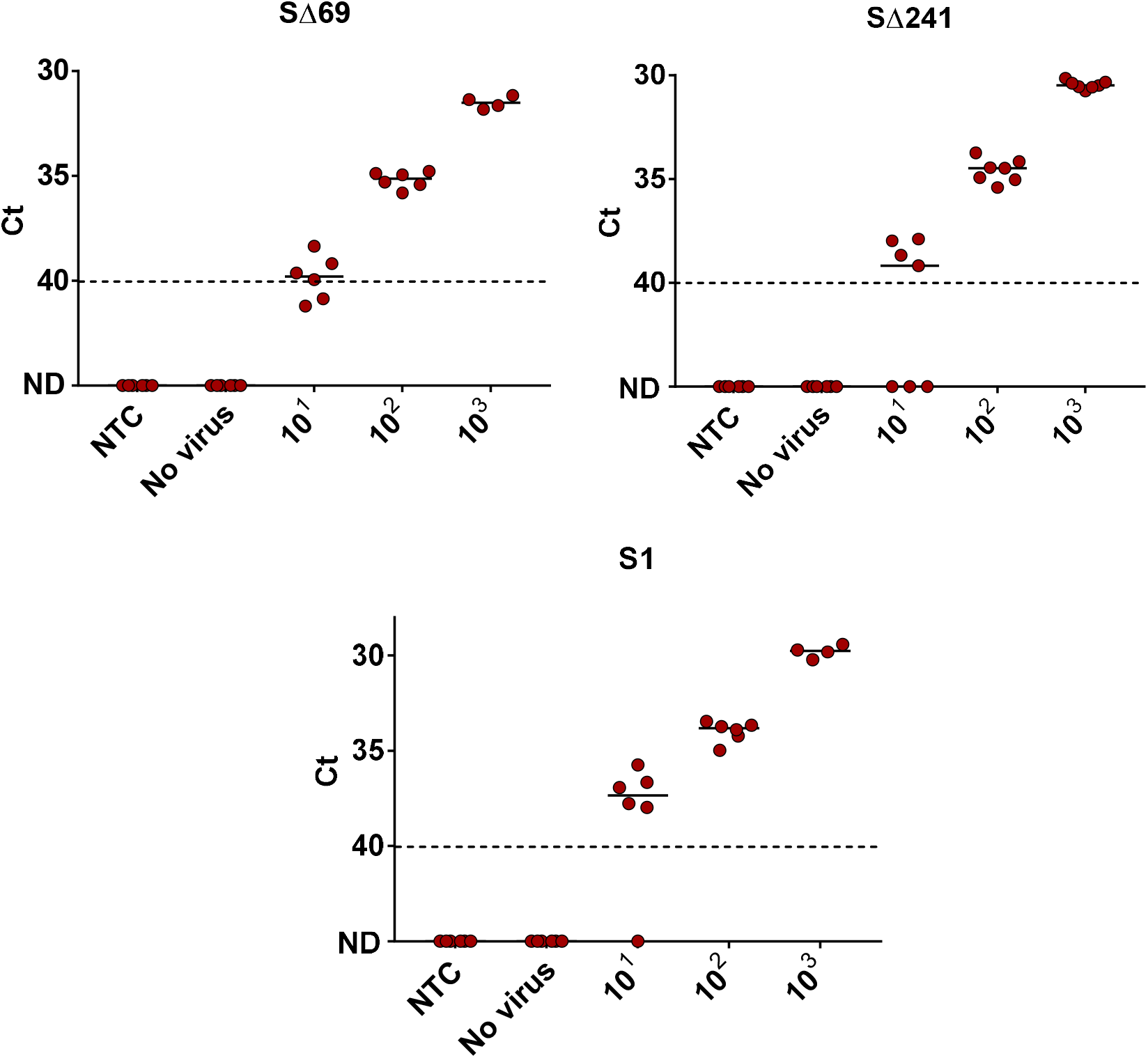
Lower detection limit of SΔ69, SΔ241 and S1 primer–probe sets in wastewater matrix. RNA extracted from negative detection wastewater sample (No virus) spiked with known concentrations of SARS-CoV-2 S gene template (10^1^–10^3^ S gene template copies?μl^−1^) and Non-Template Control (NTC, water). For SΔ69 probe and SΔ241 probe, the S gene deletion template corresponded to Δ69-70 deletion site in British B.1.1.7 and Δ241-243 deletion site in South Africa B.1.135. For S1 probe, the S gene template corresponded to NC_045512.2 sequence. ND - not detected. Solid lines indicate the median and dashed lines indicate the detection limit as decided by clinical guidelines.

To examine the probes specificity and rule out possible false-positive cases, each probe was tested with a negative control. For S1 probe, the negative control was comprised of a dsDNA template with the S gene with Δ69-70 and Δ241-243 nucleotides deletions. While for SΔ69 probe and SΔ241 probe, the original S gene sequence was used as negative control. As expected, none of the probes manifested a signal in the presence of a negative control and non-specific detection was not observed (Table 1).

Finally, the three probes, S, SΔ69 and SΔ241 were tested on wastewater samples (Table 1). Moreover, the CDC’s N2 detection set was used as standard detection reference that can correspond to each of the variants (Lu et al., 2020). According to the results, samples collected at November-December 2020 (prior to the B.1.1.7 outbreak in Israel) resulted in a positive signal from the S1 probe, while there was no signal from SΔ69 probe or SΔ241 probe. A later sample from February 2021 (B.1.1.7 was already present in Israel), resulted in a different result, as SΔ69 probe revealed positive detection, while S1 probe and SΔ241 showed no detection signal. This meant that most likely the wastewater sample from November 2020 was positive only for the original SARS-CoV-2 (NC_045512.2), while the wastewater sample from February 2021 was positive only for B.1.1.7 variant. Considering that B.1.351 variant was not detected in Israel at that time, the results matched our expectations and the SΔ241 probe did not detect any signal in any of the wastewater samples.

Looking at the detection results from wastewater from Beer-Sheva, Israel, an interesting observation was seeing that the N gene detection constantly produced lower Ct values compared to the S gene detection (with either S1 probe or SΔ69 probe). This may imply different gene expression distributions or different durability of the RNA segments, however this needs further study and validation to better understand such an observation. In the meantime, this phenomenon may also affect the “drop-out” assays resulting in false-positives, reinforcing the need in direct detection. Overall, the displayed results indicate that the developed assay can be employed and provide essential, direct detection abilities for the two variants.

**Table 3.** Detection sets results when employed on positive template, negative template or a wastewater sample.

| Sample | Sample date collected | S1 set Ct | SΔ69 set Ct | SΔ241 set Ct | CDC N2 Ct |
| --- | --- | --- | --- | --- | --- |
| 10<sup>3</sup> S gene Original template | - | 28.61 | ND** | ND** | ND** |
| 10<sup>3</sup> S gene deletion template | - | ND** | 30.29 | 29.94 | ND** |
| WWTP* Beer-Sheva | November 17, 2020 | 36.23 | ND** | ND** | 31.88 |
| WWTP* Beer-Sheva | December 8, 2020 | 35.16 | ND** | ND** | 33.89 |
| WWTP* Beer-Sheva | February 4, 2021 | ND** | 41 | ND | 33.5 |

\*WWTP-Wastewater treatment plant; \*\*ND-Not Detected

## Conclusions

The ongoing concern regarding the COVID-19 pandemic and the emergence of new variants with higher infection rate and morbidity, create great global concern. With regards to the highly dominant variants of concern, British variant B.1.1.7 and South Africa variant B.1.351, current diagnostic tools are expensive, time-consuming or indirect. Here we present an RT-qPCR assay developed for the direct detection of these two variants. An RT-qPCR assay was developed for the direct detection of the British variant (B.1.1.7) and its differentiation from the original SARS-CoV-2 (NC_045512.2). Using a single set comprised of two new primers, two new probes were designed and validated, focusing on the characterized deletion area known as Δ69-70. In addition, an RT-qPCR direct detection assay was developed for the South Africa variant (B.1.351), using two new primers focusing on a characterized deletion area known as Δ241-243, and a third probe was designed and validated. The presented primers and probe sets may be used as described here, or even combined in the future in different combinatorial approaches for rapid, cost-effective and direct detection of the two variants.

## Data Availability

All data used is available in the manuscript.

## Author Contributions

K.Y. and E.O. share equal contribution to this manuscript. K.Y. designed sequence, conceived, performed and analyzed experiments and authored this manuscript. E.O. designed sequence, conceived and analyzed experiments and authored this manuscript. N.P. and N.S.B. took part in experiments execution. A.K. conceived experiments, supervised, provided research facilities and edited the manuscript.

## Funding Sources

We would like to acknowledge funding from Ben Gurion University, The Corona Challenge Covid-19 (https://in.bgu.ac.il/en/corona-challenge/Pages/default.aspx) and funding from the Israeli ministry of Health.

## Notes

### Competing Interest Statement

The authors have declared no competing interest.

